# Emerging Applications of NLP and Large Language Models in Gastroenterology and Hepatology: A Systematic Review

**DOI:** 10.1101/2024.06.26.24309567

**Authors:** Mahmud Omar, Kassem SharIf, Benjamin Glicksberg, Girish N Nadkarni, Eyal Klang

**Affiliations:** Tel-Aviv University, Faculty of Medicine; Department of Gastroenterology, Sheba Medical Center, Tel-Hashomer, Israel; Division of Data-Driven and Digital Medicine (D3M), Icahn School of Medicine at Mount Sinai, New York, New York

**Keywords:** Natural Language Processing, Large Language Models, Gastroenterology, Hepatology, Electronic Health Records

## Abstract

**Background and Aim:** In the last two years, natural language processing (NLP) has transformed significantly with the introduction of large language models (LLM). This review updates on NLP and LLM applications and challenges in gastroenterology and hepatology.

**Methods:** Registered with PROSPERO (CRD42024542275) and adhering to PRISMA guidelines, we searched six databases for relevant studies published from 2003 to 2024, ultimately including 57 studies.

**Results:** Our review notes an increase in relevant publications in 2023-2024 compared to previous years, reflecting growing interest in newer models such as GPT-3 and GPT-4. The results demonstrate that NLP models have enhanced data extraction from electronic health records and other unstructured medical data sources. Key findings include high precision in identifying disease characteristics from unstructured reports and ongoing improvement in clinical decision-making. Risk of bias assessments using ROBINS-I, QUADAS-2, and PROBAST tools confirmed the methodological robustness of the included studies.

**Conclusion:** NLP and LLMs can enhance diagnosis and treatment in gastroenterology and hepatology. They enable extraction of data from unstructured medical records, such as endoscopy reports and patient notes, and for enhancing clinical decision-making. Despite these advancements, integrating these tools into routine practice is still challenging. Future work should prospectively demonstrate real-world value.

## WHAT IS KNOWN

- Natural language processing (NLP) models can extract data from unstructured medical records.
- Large language models (LLMs) like GPT-3 and GPT-4 are increasingly used in healthcare.
- NLP aids in identifying disease characteristics and improving clinical decision-making.

## WHAT IS NEW HERE

- This is the largest systematic review in the field of NLP and LLM applications in gastroenterology and hepatology, showing a notable increase in related publications during 2023-2024.
- NLP models show high precision in extracting data from electronic health records.
- Despite advancements, integrating these tools into routine clinical practice remains challenging.
- Future studies need to demonstrate real-world value prospectively.

## Introduction

Recent advances in Natural Language Processing (NLP) show potential for being integrated in the field of gastroenterology and hepatology (1,2). Since the last review in 2014 by Hou et al., which underscored the potential of NLP to enhance the field’s efficiency (2), there have been significant strides in technology, particularly with the emergence of Large Language Models (LLMs) such as Generative Pre-trained Transformer (GPT) and Bidirectional Encoder Representations from Transformers (BERT) (3). These developments have expanded the scope of NLP applications, from automating routine tasks to enabling complex diagnostic and therapeutic decisions (4).

NLP and LLMs extract and interpret data from patient records, notes, and reports (5–7). In gastroenterology and hepatology, they streamline the review of endoscopy, radiology, and pathology reports. This technology can help create research cohorts for clinical trials, flag complications, and support decision-making systems. Examples include managing complex conditions like IBD and hepatocellular carcinoma (5,7,8).

This review discusses the current applications and challenges of NLP and LLMs in gastroenterology and hepatology.

## Methods

### Registration and Protocol

This systematic literature review was registered with the International Prospective Register of Systematic Reviews, PROSPERO, under the registration code CRD42024542275 (9). Our methodology adhered to the Preferred Reporting Items for Systematic Reviews and Meta-Analyses (PRISMA) guidelines (10).

### Search Strategy

We conducted a rigorous search of six key databases (PubMed, Embase, Web of Science, and Scopus, Cochrane library and IEEE Xplore) for studies published until April 2024. Our focus was on the outcomes of integrating NLP and LLM models in gastroenterology and hepatology. We designed Boolean search strings tailored to each database. To maximize coverage, we supplemented our search with a comprehensive manual reference screening of included studies and targeted searches on Google Scholar. Details of the specific Boolean strings used are provided in the **Supplementary Materials**.

### Study screening and selection

Our review encompasses original research articles, and full conference papers (11). The exclusion criteria were confined to preprints, review papers, case reports, commentaries, protocol studies, editorials, and non-English publications. For the initial screening, we used the Rayyan web application (12). The initial screening and study selection, which were conducted according to predefined criteria, were independently performed by two reviewers (MO and EK).

Discrepancies were resolved through discussion. Fleiss’ kappa was calculated for the agreement between the two independent reviewers.

### Data Extraction

Data extraction was conducted by researchers MO and EK using a standardized form to ensure consistent and accurate data capture. This included details such as author, publication year, sample size, data type, task type, specific field, model used, results, numeric metrics, conclusions, and limitations. Any discrepancies in data extraction were resolved through discussion and a third reviewer was consulted when necessary.

### Risk of Bias Assessment

To ensure a thorough evaluation of the included studies, we used three tools, each tailored to a specific study design within our review. The Risk Of Bias In Non-randomized Studies of Interventions (ROBINS-I) tool has been employed in interventional studies assessing NLP in applications such as management, prescription guidance, and clinical inquiry responses (13). For diagnostic studies where NLP models were compared with physicians or a reference standard for diagnosing and detection, the Quality Assessment of Diagnostic Accuracy Studies-2 (QUADAS-2) tool was used (14). Finally, the Prediction model Risk Of Bias ASsessment Tool (PROBAST) tool was utilized for the remaining studies, which involved NLP models prediction, without direct comparison to reference standards (15). This multitool approach allowed us to appropriately address the diverse methodologies and applications presented in the reviewed studies.

## Results

### Search Results and Study Selection

A total of 720 articles were identified through initial screening. After the removal of 114 duplicates, 606 articles remained for further evaluation. Title and abstract screening led to the exclusion of 524 articles, leaving 82 articles for full-text review. Of these, the reasons for exclusion and the number of articles excluded for each reason remain the same as described earlier. Ultimately, 55 studies met all inclusion criteria. By employing reference checking and snowballing techniques, two additional studies were identified, resulting in a final tally of 57 studies (16–72). A PRISMA flowchart visually represents the screening process in **Figure 1**. Fleiss’ kappa for the agreement between screeners was calculated as 0.957, which is considered very high (73).

**Figure 1:**
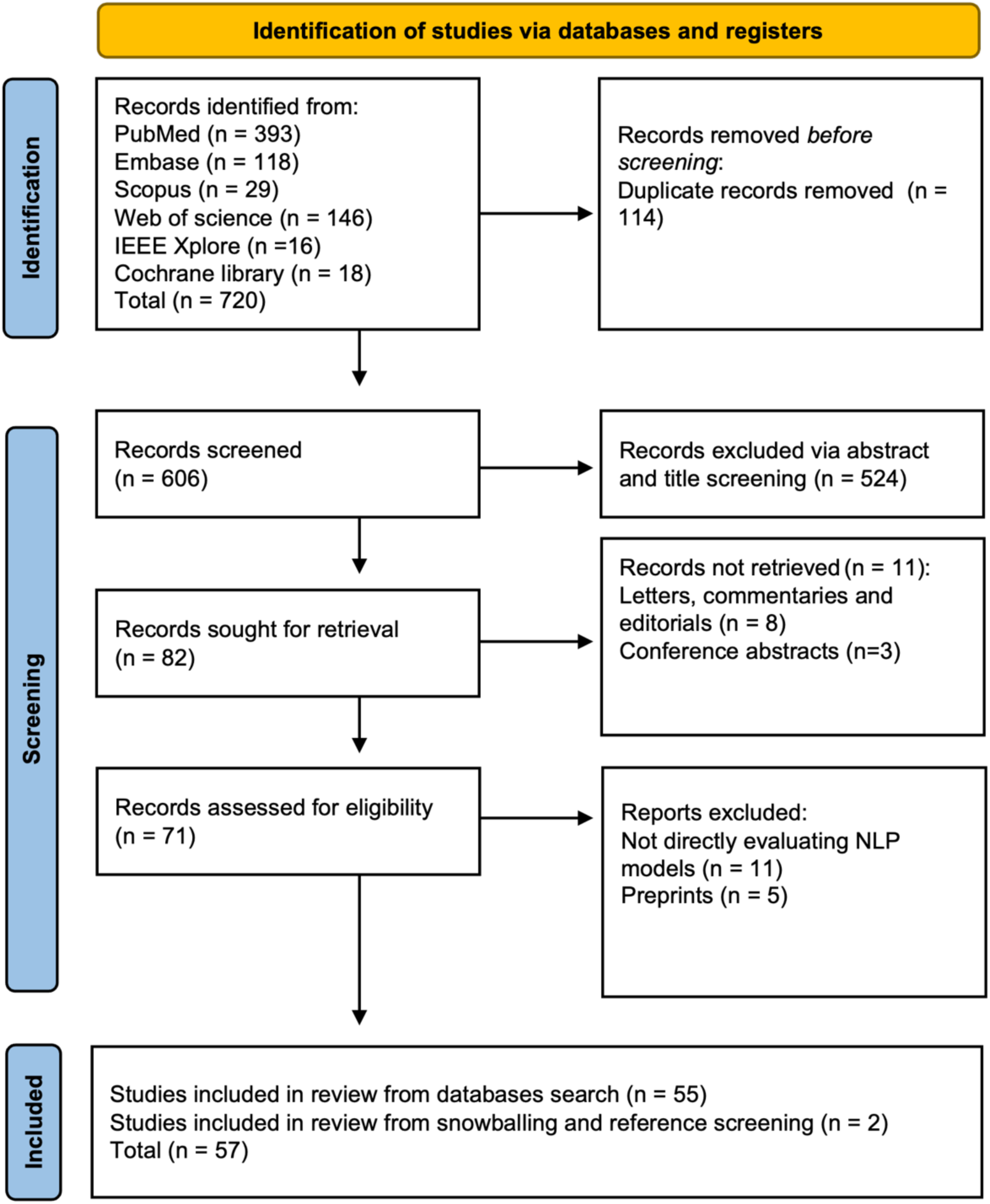
PRISMA flowchart.

### An overview of the included studies

Our systematic review incorporates a total of 57 studies (16–72). Among these, a substantial majority, 49 studies, are centered on gastroenterology, while hepatology is the focus of 8 studies. These studies span from 2018 to 2024, with a notable increase in publications in the last two years, particularly between 2023 and 2024, which collectively account for 28 of the total included studies. This uptick highlights a growing interest in advanced NLP models like GPT-3 and GPT-4.

The models employed in these studies vary widely, with traditional NLP methods and more recent LLMs like GPT-3 and GPT-4. For instance, Kong et al. (2024) utilized GPT-4 among other versions for medical counseling (38), while Schneider et al. (2023) employed rule-based NLP algorithms for detecting undiagnosed hepatic steatosis (54).

Sample sizes in these studies range from very small datasets to large-scale analyses involving millions of data points, such as in the study by Schneider et al., which analyzed data from over 2.7 million imaging reports (54). The type of data analyzed also varies significantly, encompassing electronic health records (EHRs), pathology reports, and data generated from AI models responding to preset medical queries.

Tasks performed by these models are equally diverse, from diagnostic assistance and disease monitoring to providing patient education and supporting clinical decision-making. Specific examples include the work by Truhn et al. (2024), which focused on extracting structured data from colorectal cancer reports (49), and Lahat et al. (2023), who evaluated the utility of GPT models in answering patient questions related to gastroenterology (47).

### Risk of bias

We used ROBINS-I, QUADAS-2, and PROBAST to map potential biases. Notably, most of the included studies were published in Q1 journals, affirming their scholarly impact and supported by strong SCImago Journal Rank (SJR) scores (**Figure 2**).

**Figure 2:**
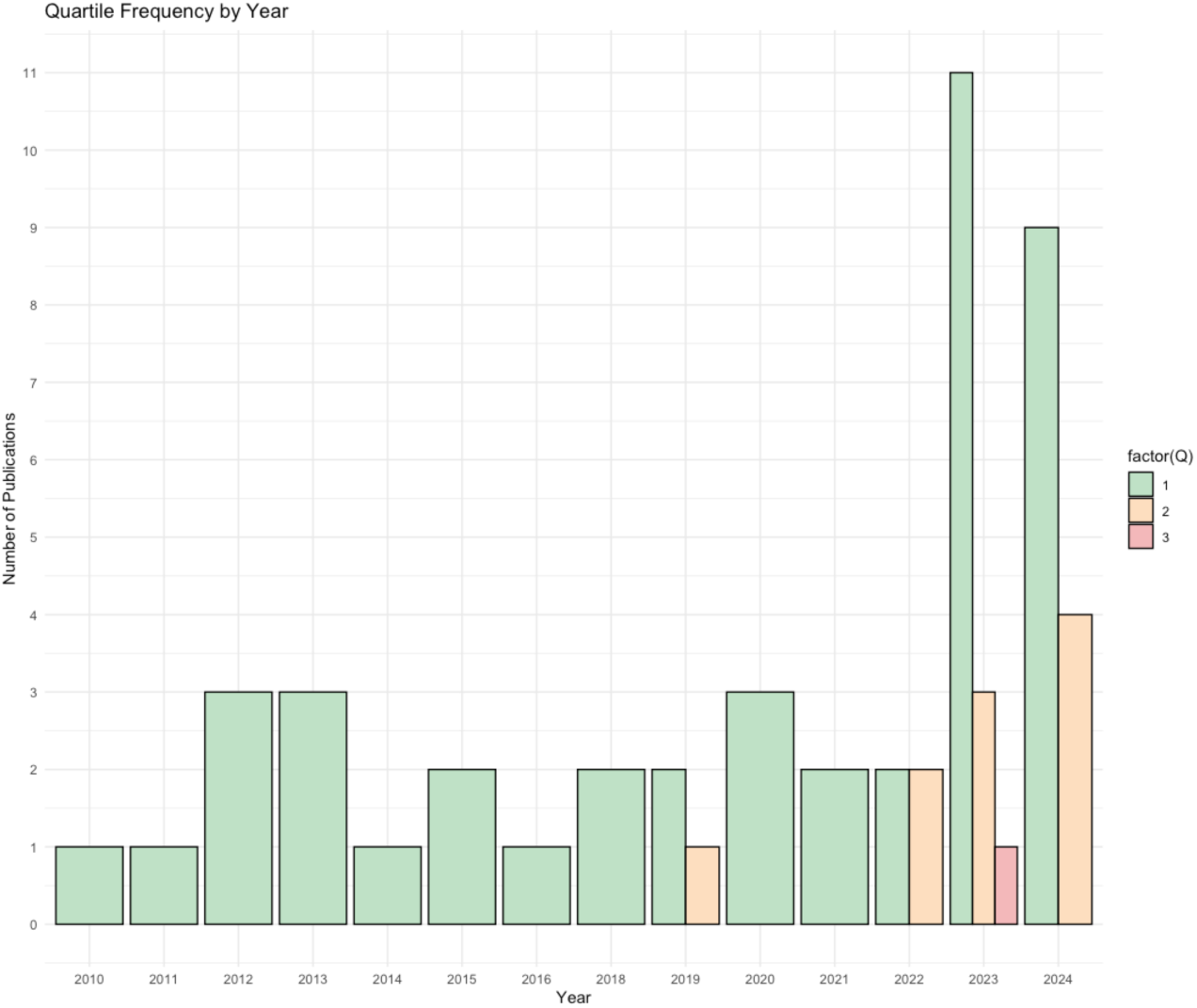
Trends of the included studies.

<u>PROBAST Results</u> (**Table S1**): This assessment mostly highlighted low-risk ratings in outcome and analysis domains. However, several studies encountered issues with high participant-related applicability biases, influencing the generalizability of their findings.

<u>QUADAS-2 Results</u> (**Table S2**): A synthesis of QUADAS-2 results revealed that most studies (20 out of 32) exhibited low risk of bias across all four assessed domains. This underscores their methodological robustness and reliability. However, three studies were identified as having a high risk of bias in one of the four categories. Patient selection applicability concerns were notable, primarily due to the reliance on single-center data with specific documentation styles, which may limit the broader applicability of these findings.

<u>ROBINS-I Results</u> (**Table S3**): Analysis of ROBINS-I revealed that 14 studies displayed a moderate risk of bias overall, while one study exhibited a high risk. This was largely due to biases in the selection of participants into the study and confounding factors, particularly because many studies utilized specific questions, queries, or fictional vignettes and case scenarios. Despite these concerns, the other assessment categories predominantly showed low risk. Nonetheless, six studies demonstrated low risk across all evaluated domains.

### NLP Applications

We categorized the applications of the NLP and LLM models under three main categories for a synthesized analysis of the results: Disease Detection and Diagnosis (n = 30), Patient Care (n = 22), and Education and Research applications (n = 5). Disease Detection and Diagnosis was further divided into Colonoscopy Reports and Other Diagnostic Applications, while Patient Care included Management and Communication and Clinical Decision Support, focusing on patient-oriented and healthcare professionals-oriented applications, respectively (**Figure 3**).

**Figure 3:**
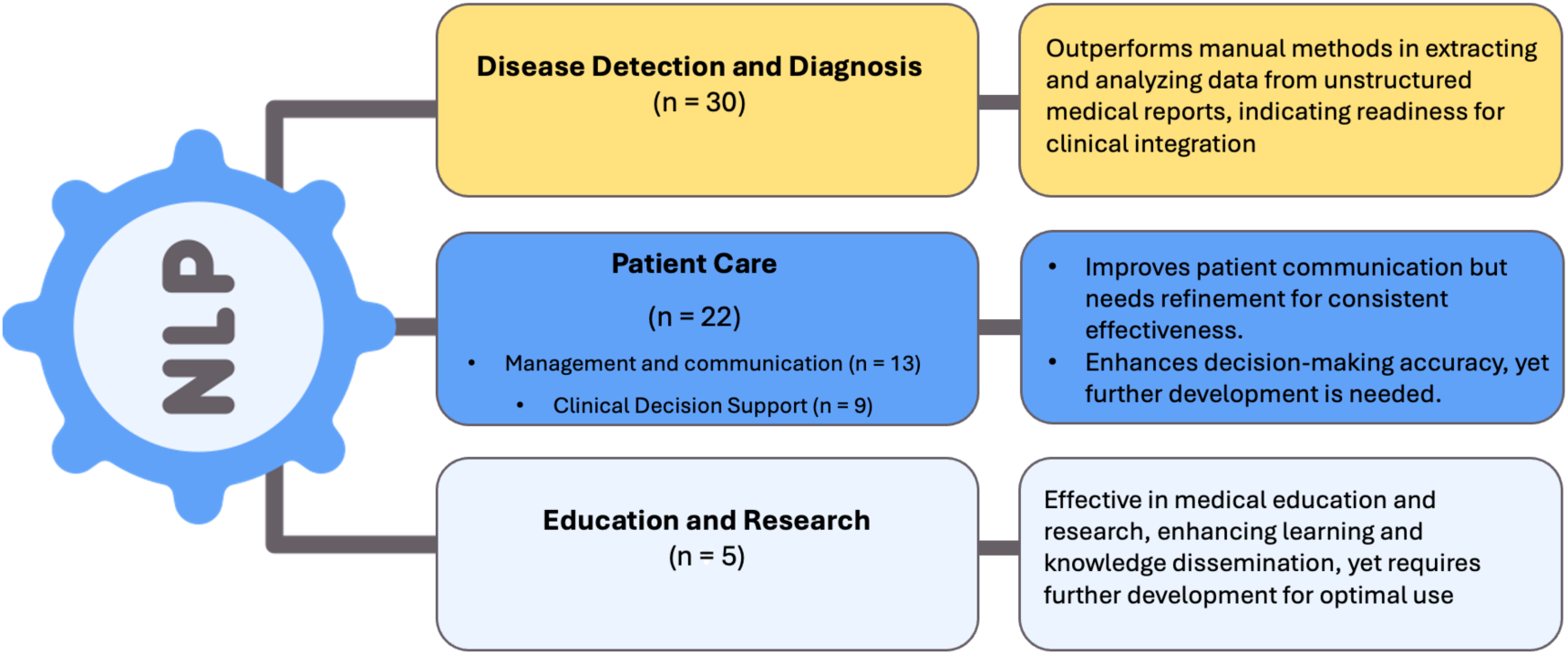
Summary of NLP applications and outcomes.

### Disease Detection and Diagnosis

Most of the studies evaluated NLP models in extracting data from unstructured colonoscopy reports (n=17) (**Figure 4**). Nonetheless, there were many unique applications.

**Figure 4:**
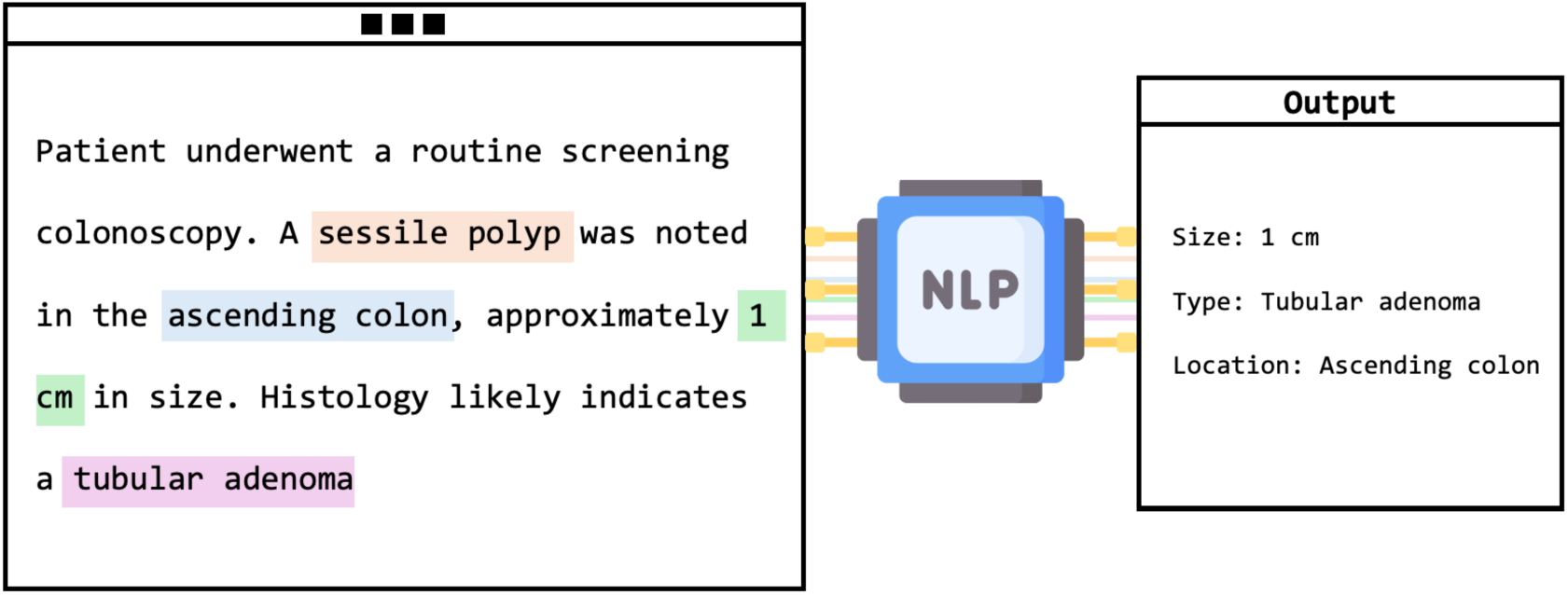
Visual framework of NLP extracting adenoma characteristics from unstructured colonoscopy report.

<u>Colonoscopy Reports:</u> This category, which includes 17 studies, primarily explored NLP’s role in enhancing the interpretation of unstructured colonoscopy reports. Various quality and diagnostic measures were evaluated, such as the adenoma detection rate (ADR), a frequent subject of investigation. For instance, Nayor et al. reported that their NLP pipeline achieved high precision and recall in the automated calculation of ADR (57). Other assessments included polyp detection and sizing, with Imler et al. demonstrating accuracies of 98% for pathology level identification and 96% for size estimation (58). Additionally, Raju et al. noted that NLP matched or exceeded manual methods in identifying and categorizing adenomas with a detection rate of 43% (59). Overall, NLP models showed a broad range of accuracies from 84% to 100%, consistently outperforming manual review methods. Despite needing GPUs, these models reduce the time and effort of manual evaluations.

Other Diagnostic Applications: Beyond colonoscopy, NLP was applied to a diverse array of diagnostic contexts in gastroenterology and hepatology.

In gastroenterology, several innovative NLP applications have emerged. For example, Wenker et al. utilized NLP to identify dysplasia in Barrett’s Esophagus from esophagogastroduodenoscopy (EGD) reports with a high accuracy of 98.7% (69). Song et al. developed a model to extract detailed clinical information such as disease presence, location, size, and stage from unstructured EGD reports, achieving high sensitivity, precision, and accuracy scores (61). Denny et al. applied NLP to enhance colorectal cancer screening by identifying references to four CRC tests within electronic clinical documentation, demonstrating superior recall compared to traditional manual and billing record reviews (63). Additionally, Blumenthal et al. and Parthasarathy et al. used NLP for patient monitoring, with the former detecting non-adherence to follow-up colonoscopies with an AUC of 70.2%, and the latter identifying patients meeting WHO criteria for serrated polyposis syndrome with 93% accuracy (18,65).

For IBDs, Stidham et al. utilized NLP to detect and infer the activity status of extraintestinal manifestations from clinical notes, enhancing detection accuracy to 94.1% and specificity to 95% (52). Ananthakrishnan et al. explored improving case definitions for Crohn’s disease and ulcerative colitis by combining codified data with narrative clinical texts, which identified 6-12% more patients than models using codified data alone, with AUCs of 0.95 for Crohn’s disease and 0.94 for ulcerative colitis (53).

In hepatology, NLP has facilitated significant advancements in disease identification and progression monitoring. Sada et al. combined NLP with ICD-9 codes to improve the identification of hepatocellular carcinoma cases from EHR data, significantly enhancing sensitivity and specificity, with an F2 score of 0.92 (71). Van Vleck et al. employed NLP to track disease progression in patients with non-alcoholic fatty liver disease (NAFLD), demonstrating superior sensitivity and F2 scores compared to traditional methods, effectively identifying disease progression from NAFLD to NASH or cirrhosis with sensitivity of 0.93 and an F2 score of 0.92 (30). Furthermore, Sherman et al. developed an NLP model capable of automatically scoring and classifying histological features found in pathology reports related to metabolic associated steatohepatitis (17). The goal was to estimate the risk of progression towards cirrhosis. The model demonstrated high positive and negative predictive values, ranging from 93.5% to 100%, across various histological features (17). Importantly, this NLP model facilitated the creation of a large and quality-controlled cohort of MASLD patients (17).

### Patient Care

The patient care section is subdivided into two categories: patient management and communication, which comprises 13 studies, and clinical decision support, encompassing 9 studies.

Management and Communication: This category explores the use of NLP and LLMs in facilitating communication and management.

In gastroenterology, studies like Lahat et al. evaluated ChatGPT’s ability to answer real-life gastroenterology-related patient queries, achieving moderate effectiveness with accuracy scores ranging from 3.4 to 3.9 (47). Choo et al. reported an 86.7% concordance rate between ChatGPT’s recommendations for managing complex colorectal cancer cases and decisions made by multidisciplinary teams (39). Furthermore, Lim et al. demonstrated that a contextualized GPT-4 model provided accurate colonoscopy interval advice, significantly outperforming standard models by adhering closely to established guidelines (33). Imler et al. used the cTAKES system to achieve an 81.7% agreement with guideline-adherent colonoscopy surveillance intervals, substantially surpassing manual review accuracies (36). However, studies like Huo et al. and Atarere et al. indicated variability in ChatGPT’s performance, suggesting the need for enhancements in AI consistency and reliability (25,44). In the area of IBD, Zand et al. developed an NLP model that categorized electronic dialog data, showing a 95% agreement with physician evaluations and underscoring the potential of automated chatbots in patient interaction (23). Sciberras et al. found ChatGPT to provide highly accurate (84.2%) and moderately complete responses to patient inquiries about IBD, with particular strengths in topics like smoking and medication (20).

In hepatology, Yeo et al. tested GPT’s proficiency in delivering emotional support and accurate information on cirrhosis and hepatocellular carcinoma, achieving correct response rates of 79.1% for cirrhosis and 74% for carcinoma (29). Samaan et al. explored GPT’s effectiveness in Arabic, noting a 72.5% accuracy rate, though it was less accurate than its English counterpart, indicating disparities in language performance (34).

<u>Clinical Decision Support</u>: NLP models were tested for their accuracy and effectiveness in decision-making scenarios. For example, Kong et al. evaluated LLMs’ capability to provide counseling on Helicobacter pylori, noting that while accuracy was generally high (90% acceptable responses), completeness needed improvement (38). Li et al.’s integration of NLP with machine learning for predicting liver metastases showed impressive results with accuracy and F1 scores around 80.4% (24). The study by Becker et al. utilized an NLP pipeline tailored for German, achieving high precision and recall in guideline-based treatment extraction from clinical notes (64). Further, Wang et al.’s “DeepCausality” framework accurately assessed causal factors for drug-induced liver injuries, aligning well with clinical guidelines (41). Another significant study, Wagholikar et al., demonstrated that an NLP-powered clinical decision support system could assist in making guideline-adherent recommendations for colonoscopy surveillance, as it made optimal recommendations in 48 out of 53 cases (35).

### Education and Research

Five studies focused on this aspect. Generaly, NLP and LLMs have demonstrated a promising capacity to enhance learning and knowledge dissemination. Benedicenti et al. explored the accuracy of ChatGPT in solving clinical vignettes against gastroenterologists, noting an initial 40% accuracy that improved to 65% over time, suggesting a potential for future clinical integration with continued advancements (56). Zhou et al. assessed GPT-3.5 and GPT-4 for their ability to provide consultation recommendations and analyze gastroscopy reports related to gastric cancer, with GPT-4 achieving 91.3% appropriateness and 95.7% consistency (48). Lahat et al. utilized GPT to generate research questions in gastroenterology, finding the questions relevant and clear but lacking in originality (46). Meanwhile, Gravina et al. highlighted the efficacy of ChatGPT 3.5 in medical education, as it outperformed Perplexity AI in residency exam questions with a 94.11% accuracy rate (32). Additionally, Pradhan et al. compared AI-generated patient educational materials on cirrhosis with human-derived content, finding no significant differences in readability or accuracy, though human materials were deemed more actionable (28).

## Discussion

Our systematic review assessed the integration of NLP and LLMS in gastroenterology and hepatology, registering significant advancements. We reviewed 57 studies, highlighting a sharp increase in research over the last two years, particularly focusing on newer models like GPT-3 and GPT-4. These studies reflect a shift from traditional tasks, such as report analysis, to more dynamic roles in patient management and research facilitation.

The results show that certain NLP applications seem ready for immediate clinical use. For example, Schneider et al. (2023) identified 42,000 hepatic steatosis cases using an NLP model on 2.15 million pathology reports and 2.7 million imaging reports. This level of precision (PPV 99.7%) exemplifies NLP’s readiness to support diagnostic processes in large-scale healthcare settings. Similarly, Truhn et al. (2024) successfully employed GPT-4 to extract structured data from colorectal cancer reports with a precision of 99% for T-stage identification, suggesting a high reliability of NLP in processing and structuring complex pathological data.

Conversely, the technology’s expansion into more dynamic roles such as comprehensive disease management and holistic patient care is still evolving. For instance, Kong et al. (2024) found that while the accuracy and comprehensibility of GPT-4’s responses to medical inquiries about Helicobacter pylori were high, the completeness of the information was less satisfactory. This indicates ongoing challenges in ensuring that NLP outputs are not only accurate but also fully informative.

Our results suggest that both classic NLP methods and newer models can be effectively integrated to streamline manual tasks such as extracting data and making diagnoses from complex and unstructured reports, with an accuracy that typically surpasses manual screening (16–18,21,22,27,33). This builds upon and adds on a previous systematic review of NLP in gastroenterology and hepatology conducted by Hou et al. (2). While he found promising results, he emphasized the need for careful consideration of the quality of clinical data within EHRs, and also highlighted the importance of understanding variations and deviations from established clinical practice standards (2). Our updated and more comprehensive results indicate that these models consistently demonstrate high accuracies (16–18,21,22,27,33). This trend is observable in other fields utilizing NLP, such as radiology and infectious diseases (74,75). However, our research suggests that applying these methods to more complex tasks like patient management, education, and clinical decision-making is still challenging (20,29,34,37). While newer models show promising results, there are significant limitations and variability that require further development (67). This trend is consistent with data and the current findings from other fields (76,77).

Several limitations of our review must be acknowledged. Many studies utilize single-institution datasets, which could affect the generalizability of the findings. The accuracy of NLP outputs is heavily dependent on the quality of the input data, with errors or inconsistencies in medical records potentially leading to inaccurate results (78). The opaque nature of AI decision-making processes (’black box’) raises concerns about the transparency and trustworthiness of these models in clinical settings (79). Finally, ethical considerations around potential biases in training data and algorithmic outputs underscore the necessity for careful implementation to ensure fairness and equity in healthcare delivery (80).

In conclusion,

our systematic review highlights the impact of NLP and LLMs in gastroenterology and hepatology. On one hand, NLP has already proven its utility in screening and analyzing medical reports, facilitating streamlined screening policies with impressive outcomes. On the other hand, the capabilities of newer LLMs are still unfolding, with their full potential in complex management and research roles yet to be fully realized.

The results demonstrate that while some applications of NLP are well-established and highly effective, newer LLMs offer exciting, emerging applications that promise to further enhance clinical practice. Moving forward, research focus should be on refining these models to ensure prospectively they meet real-world clinical needs.

## Data Availability

All data produced in the present study are available upon reasonable request to the authors

## Acknowledgment

none.

## Competing interest

None declared.

**Ethical approval** was not required for this research.

## Data Sharing Statement

This manuscript is a systematic review; therefore, all data utilized are available in the published articles that were reviewed. As this study did not generate new data, but rather analyzed existing published data, there are no additional datasets to share.

## Contributorship

- M.O: Responsible for writing the initial draft and the final version of the manuscript, gathering and extracting data, and visualizing the results.
- K.S: Provided oversight and editing of the manuscript.
- B.G: Provided oversight and editing of the manuscript.
- G.N: Oversaw the entire project.
- E.K: Involved in data extraction, editing the final manuscript, and validation of the data.

## Specific Booleans used in each database

### PubMed

| Search number | Query | Filters | Search Details | Results |
| --- | --- | --- | --- | --- |
| 5 | ("natural language processing" OR "NLP" OR "large language models" OR "LLM" OR "BERT" OR "GPT-3" OR "GPT" OR "ChatGPT" OR "GPT-3.5" OR "GPT-4" OR "transformer language models") AND ("gastroenterology" OR "digestive health" OR "gastrointestinal oncology" OR "endoscopic imaging" OR "colorectal cancer screening" OR "hepatocellular carcinoma" OR "liver fibrosis" OR "inflammatory bowel disease management" OR "IBD" OR "Crohn's disease" OR "ulcerative colitis treatment") | English, from 2003 - 2024 | ((("natural language processing"[All Fields] OR "NLP"[All Fields] OR "large language models"[All Fields] OR "LLM"[All Fields] OR "BERT"[All Fields] OR "GPT-3"[All Fields] OR "GPT"[All Fields] OR "ChatGPT"[All Fields] OR "GPT-3.5"[All Fields] OR "GPT-4"[All Fields] OR "transformer language models"[All Fields]) AND ("gastroenterology"[All Fields] OR "digestive health"[All Fields] OR "gastrointestinal oncology"[All Fields] OR "endoscopic imaging"[All Fields] OR "colorectal cancer screening"[All Fields] OR "hepatocellular carcinoma"[All Fields] OR "liver fibrosis"[All Fields] OR "inflammatory bowel disease management"[All Fields] OR "IBD"[All Fields] OR "Crohn's disease"[All Fields] OR "ulcerative colitis treatment"[All Fields])) AND ((english[Filter]) AND (2003:2024[pdat])) | 393 |
| 4 | ("natural language processing" OR "NLP" OR "large language models" OR "LLM" OR "BERT" OR "GPT-3" OR "GPT" OR "ChatGPT" OR "GPT-3.5" OR "GPT-4" OR "transformer language models") AND ("gastroenterology" OR "digestive health" OR "gastrointestinal oncology" OR "endoscopic imaging" OR "colorectal cancer screening" OR "hepatocellular carcinoma" OR "liver fibrosis" OR "inflammatory bowel disease management" OR "IBD" OR | from 2003 - 2024 | ((("natural language processing"[All Fields] OR "NLP"[All Fields] OR "large language models"[All Fields] OR "LLM"[All Fields] OR "BERT"[All Fields] OR "GPT-3"[All Fields] OR "GPT"[All Fields] OR "ChatGPT"[All Fields] OR "GPT-3.5"[All Fields] OR "GPT-4"[All Fields] OR "transformer language models"[All Fields]) AND ("gastroenterology"[All Fields] OR "digestive health"[All Fields] OR "gastrointestinal oncology"[All Fields] OR "endoscopic imaging"[All Fields] OR "colorectal cancer screening"[All Fields] OR "hepatocellular carcinoma"[All Fields] OR "liver fibrosis"[All Fields] OR "inflammatory bowel disease management"[All Fields] OR "IBD"[All Fields] OR "Crohn's disease"[All | 404 |
|  | "Crohn's disease" OR "ulcerative colitis treatment") |  | Fields] OR "ulcerative colitis treatment"[All Fields])) AND (2003:2024[pdat]) |  |
| 3 | ("natural language processing" OR "NLP" OR "large language models" OR "LLM" OR "BERT" OR "GPT-3" OR "GPT" OR "ChatGPT" OR "GPT-3.5" OR "GPT-4" OR "transformer language models") AND ("gastroenterology" OR "digestive health" OR "gastrointestinal oncology" OR "endoscopic imaging" OR "colorectal cancer screening" OR "hepatocellular carcinoma" OR "liver fibrosis" OR "inflammatory bowel disease management" OR "IBD" OR "Crohn's disease" OR "ulcerative colitis treatment") |  | ("natural language processing"[All Fields] OR "NLP"[All Fields] OR "large language models"[All Fields] OR "LLM"[All Fields] OR "BERT"[All Fields] OR "GPT-3"[All Fields] OR "GPT"[All Fields] OR "ChatGPT"[All Fields] OR "GPT-3.5"[All Fields] OR "GPT-4"[All Fields] OR "transformer language models"[All Fields]) AND ("gastroenterology"[All Fields] OR "digestive health"[All Fields] OR "gastrointestinal oncology"[All Fields] OR "endoscopic imaging"[All Fields] OR "colorectal cancer screening"[All Fields] OR "hepatocellular carcinoma"[All Fields] OR "liver fibrosis"[All Fields] OR "inflammatory bowel disease management"[All Fields] OR "IBD"[All Fields] OR "Crohn's disease"[All Fields] OR "ulcerative colitis treatment"[All Fields]) | 474 |
| 2 | ((("natural language processing" OR "NLP" OR "large language models" OR "LLM" OR "BERT" OR "GPT-3" OR "GPT" OR "ChatGPT" OR "GPT-3.5" OR "GPT-4" OR "transformer language models" OR "deep learning" OR "machine learning" OR "AI" OR "artificial intelligence" OR "text mining" OR "computational linguistics") AND ("gastroenterology" OR "digestive health" OR "gastrointestinal oncology" OR "endoscopic imaging" OR "colorectal cancer screening" OR "hepatocellular carcinoma" OR "liver fibrosis" OR "liver diseases" OR "gastrointestinal diseases" OR "digestive system diseases" OR | from 2003 - 2024 | ((("natural language processing"[All Fields] OR "NLP"[All Fields] OR "large language models"[All Fields] OR "LLM"[All Fields] OR "BERT"[All Fields] OR "GPT-3"[All Fields] OR "GPT"[All Fields] OR "ChatGPT"[All Fields] OR "GPT-3.5"[All Fields] OR "GPT-4"[All Fields] OR "transformer language models"[All Fields] OR "deep learning"[All Fields] OR "machine learning"[All Fields] OR "AI"[All Fields] OR "artificial intelligence"[All Fields] OR "text mining"[All Fields] OR "computational linguistics"[All Fields]) AND ("gastroenterology"[All Fields] OR "digestive health"[All Fields] OR "gastrointestinal oncology"[All Fields] OR "endoscopic imaging"[All Fields] OR "colorectal cancer screening"[All Fields] OR "hepatocellular carcinoma"[All Fields] OR "liver fibrosis"[All Fields] OR "liver diseases"[All Fields] OR "gastrointestinal diseases"[All Fields] OR "digestive system diseases"[All Fields] OR "inflammatory bowel disease management"[All Fields] OR "IBD"[All Fields] | 13,056 |
|  | "inflammatory bowel disease management" OR "IBD" OR "Crohn's disease" OR "ulcerative colitis treatment" OR "gastrointestinal inflammation" OR "gastrointestinal imaging" OR "gastrointestinal diagnostics")) |  | OR "Crohn's disease"[All Fields] OR "ulcerative colitis treatment"[All Fields] OR "gastrointestinal inflammation"[All Fields] OR "gastrointestinal imaging"[All Fields] OR "gastrointestinal diagnostics"[All Fields])) AND (2003:2024[pdat]) |  |
| 1 | ((("natural language processing" OR "NLP" OR "large language models" OR "LLM" OR "BERT" OR "GPT-3" OR "GPT" OR "ChatGPT" OR "GPT-3.5" OR "GPT-4" OR "transformer language models" OR "deep learning" OR "machine learning" OR "AI" OR "artificial intelligence" OR "text mining" OR "computational linguistics") AND ("gastroenterology" OR "digestive health" OR "gastrointestinal oncology" OR "endoscopic imaging" OR "colorectal cancer screening" OR "hepatocellular carcinoma" OR "liver fibrosis" OR "liver diseases" OR "gastrointestinal diseases" OR "digestive system diseases" OR "inflammatory bowel disease management" OR "IBD" OR "Crohn's disease" OR "ulcerative colitis treatment" OR "gastrointestinal inflammation" OR "gastrointestinal imaging" OR "gastrointestinal diagnostics")) |  | ("natural language processing"[All Fields] OR "NLP"[All Fields] OR "large language models"[All Fields] OR "LLM"[All Fields] OR "BERT"[All Fields] OR "GPT-3"[All Fields] OR "GPT"[All Fields] OR "ChatGPT"[All Fields] OR "GPT-3.5"[All Fields] OR "GPT-4"[All Fields] OR "transformer language models"[All Fields] OR "deep learning"[All Fields] OR "machine learning"[All Fields] OR "AI"[All Fields] OR "artificial intelligence"[All Fields] OR "text mining"[All Fields] OR "computational linguistics"[All Fields]) AND ("gastroenterology"[All Fields] OR "digestive health"[All Fields] OR "gastrointestinal oncology"[All Fields] OR "endoscopic imaging"[All Fields] OR "colorectal cancer screening"[All Fields] OR "hepatocellular carcinoma"[All Fields] OR "liver fibrosis"[All Fields] OR "liver diseases"[All Fields] OR "gastrointestinal diseases"[All Fields] OR "digestive system diseases"[All Fields] OR "inflammatory bowel disease management"[All Fields] OR "IBD"[All Fields] OR "Crohn's disease"[All Fields] OR "ulcerative colitis treatment"[All Fields] OR "gastrointestinal inflammation"[All Fields] OR "gastrointestinal imaging"[All Fields] OR "gastrointestinal diagnostics"[All Fields]) | 14,478 |

### Embase

(’natural language processing’ OR ‘nlp’ OR ‘bert’ OR ‘gpt-3’ OR ‘gpt’ OR ‘chatgpt’ OR ‘gpt-3.5’ OR ‘gpt-4’ OR ‘transformer language models’ OR ‘ai language technology’) AND (’gastroenterology’ OR ‘hepatology’ OR ‘gastrointestinal oncology’ OR ‘endoscopic imaging’ OR ‘colorectal cancer screening’ OR ‘hepatocellular carcinoma’ OR ‘liver fibrosis’ OR ‘inflammatory bowel disease management’ OR ‘ibd’ OR ‘crohns disease’ OR ‘ulcerative colitis treatment’)

AND (2003:py OR 2004:py OR 2005:py OR 2006:py OR 2007:py OR 2008:py OR 2009:py OR 2010:py OR 2011:py OR 2012:py OR 2013:py OR 2014:py OR 2015:py OR 2016:py OR 2017:py OR 2018:py OR 2019:py OR 2020:py OR 2021:py OR 2022:py OR 2023:py OR 2024:py) AND [embase]/lim NOT ([embase]/lim AND [medline]/lim) AND (’article’/it OR ‘conference paper’/it)

### Scopus

(TITLE-ABS-KEY((”natural language processing” OR “NLP” OR “large language models” OR “LLM” OR “BERT” OR “GPT-3” OR “GPT” OR “ChatGPT” OR “GPT-3.5” OR “GPT-4” OR “transformer language models”) AND (”Gastroenterology” OR “Hepatology” OR “peptic ulcer disease” OR “PUD” OR “Endoscopy” OR “colorectal cancer” OR “hepatocellular carcinoma treatment” OR “IBD predictive modeling” OR “endoscopic imaging”))) AND PUBYEAR > 2002 AND (NOT DOCTYPE(re) OR DOCTYPE(le) OR DOCTYPE(ed) OR DOCTYPE(cr) OR DOCTYPE(cp))

### Web of science

TS=(”natural language processing” OR “NLP” OR “large language models” OR) “LLM” OR “BERT” OR “GPT-3” OR “GPT” OR “ChatGPT” OR “GPT-3.5” OR “GPT-4” OR “transformer language models”) AND TS=(”Gastroenterology” OR “Hepatology” OR “peptic ulcer disease” OR “PUD” OR “Endoscopy” OR “colorectal cancer” OR “hepatocellular carcinoma treatment” OR “IBD predictive modeling” OR “endoscopic imaging”)) AND PY=(2003-2023) NOT DT=(”Review” “(OR “Editorial Material” OR “Letter” OR “Meeting Abstract” OR “News Item”)

### Cochrane library

(”natural language processing” OR “NLP” OR “large language models” OR “LLM” OR “BERT” OR “GPT-3” OR “GPT” OR “ChatGPT” OR “GPT-3.5” OR “GPT-4” OR “transformer language models”) AND (”gastroenterology” OR “hepatology” OR “digestive health” OR “gastrointestinal oncology” OR “endoscopic imaging” OR “colorectal cancer screening” OR “hepatocellular carcinoma” OR “liver fibrosis” OR “inflammatory bowel disease management” OR “IBD” OR “Crohn’s disease” OR “ulcerative colitis treatment”)

### IEEE Xplore

(”natural language processing” OR “NLP” OR “large language models” OR “LLM” OR “BERT” OR “GPT-3” OR “GPT” OR “ChatGPT” OR “GPT-3.5” OR “GPT-4” OR “transformer language models”) AND (”gastroenterology” OR “hepatology” OR “digestive health” OR “gastrointestinal oncology” OR “endoscopic imaging” OR “colorectal cancer screening” OR “hepatocellular carcinoma” OR “liver fibrosis” OR “inflammatory bowel disease management” OR “IBD” OR “Crohn’s disease” OR “ulcerative colitis treatment”)

## References

1. Klang E, Sourosh A, Nadkarni GN, Sharif K, Lahat A. Evaluating the role of ChatGPT in gastroenterology: a comprehensive systematic review of applications, benefits, and limitations. Ther Adv Gastroenterol. 2023;16:17562848231218618.

2. Hou JK, Imler TD, Imperiale TF. Current and future applications of natural language processing in the field of digestive diseases. Clin Gastroenterol Hepatol Off Clin Pract J Am Gastroenterol Assoc. 2014 Aug;12(8):1257–61.

3. Dave T, Athaluri SA, Singh S. ChatGPT in medicine: an overview of its applications, advantages, limitations, future prospects, and ethical considerations. Front Artif Intell. 2023;6:1169595.

4. Nehme F, Feldman K. Evolving Role and Future Directions of Natural Language Processing in Gastroenterology. Dig Dis Sci. 2021 Jan;66(1):29–40.

5. Tavanapong W, Oh J, Riegler MA, Khaleel M, Mittal B, de Groen PC. Artificial Intelligence for Colonoscopy: Past, Present, and Future. IEEE J Biomed Health Inform. 2022 Aug;26(8):3950–65.

6. Stidham RW. Artificial Intelligence for Understanding Imaging, Text, and Data in Gastroenterology. Gastroenterol Hepatol. 2020 Jul;16(7):341–9.

7. Zaver HB, Patel T. Opportunities for the use of large language models in hepatology. Clin Liver Dis. 2023 Sep 13;22(5):171–6.

8. Shahab O, El Kurdi B, Shaukat A, Nadkarni G, Soroush A. Large language models: a primer and gastroenterology applications. Ther Adv Gastroenterol. 2024;17:17562848241227031.

9. Schiavo JH. PROSPERO: An International Register of Systematic Review Protocols. Med Ref Serv Q. 2019;38(2):171–80.

10. Page MJ, McKenzie JE, Bossuyt PM, Boutron I, Hoffmann TC, Mulrow CD, et al. The PRISMA 2020 statement: an updated guideline for reporting systematic reviews. BMJ. 2021 Mar 29;372:n71.

11. Brietzke E, Gomes FA, Gerchman F, Freire RCR. Should systematic reviews and meta-analyses include data from preprints? Trends Psychiatry Psychother. 45:e20210324.

12. Ouzzani M, Hammady H, Fedorowicz Z, Elmagarmid A. Rayyan-a web and mobile app for systematic reviews. Syst Rev. 2016 Dec 5;5(1):210.

13. Sterne JA, Hernán MA, Reeves BC, Savović J, Berkman ND, Viswanathan M, et al. ROBINS-I: a tool for assessing risk of bias in non-randomised studies of interventions. BMJ. 2016 Oct 12;355:i4919.

14. Whiting PF, Rutjes AWS, Westwood ME, Mallett S, Deeks JJ, Reitsma JB, et al. QUADAS-2: a revised tool for the quality assessment of diagnostic accuracy studies. Ann Intern Med. 2011 Oct 18;155(8):529–36.

15. Wolff RF, Moons KGM, Riley RD, Whiting PF, Westwood M, Collins GS, et al. PROBAST: A Tool to Assess the Risk of Bias and Applicability of Prediction Model Studies. Ann Intern Med. 2019 Jan 1;170(1):51–8.

16. Li J, Hu S, Shi C, Dong Z, Pan J, Ai Y, et al. A deep learning and natural language processing-based system for automatic identification and surveillance of high-risk patients undergoing upper endoscopy: A multicenter study. EClinicalMedicine. 2022 Nov;53:101704.

17. Sherman MS, Challa PK, Przybyszewski EM, Wilechansky RM, Uche-Anya EN, Ott AT, et al. A natural language processing algorithm accurately classifies steatotic liver disease pathology to estimate the risk of cirrhosis. Hepatol Commun. 2024 Apr 1;8(4):e0403.

18. Parthasarathy G, Lopez R, McMichael J, Burke CA. A natural language-based tool for diagnosis of serrated polyposis syndrome. Gastrointest Endosc. 2020 Oct;92(4):886–90.

19. A Transparent and Adaptable Method to Extract Colonoscopy and Pathology Data Using Natural Language Processing - PubMed [Internet]. [cited 2024 May 1]. Available from: https://pubmed.ncbi.nlm.nih.gov/32737597/

20. Sciberras M, Farrugia Y, Gordon H, Furfaro F, Allocca M, Torres J, et al. Accuracy of Information given by ChatGPT for patients with Inflammatory Bowel Disease in relation to ECCO Guidelines. J Crohns Colitis. 2024 Mar 23;jjae040.

21. Lee JK, Jensen CD, Levin TR, Zauber AG, Doubeni CA, Zhao WK, et al. Accurate Identification of Colonoscopy Quality and Polyp Findings Using Natural Language Processing. J Clin Gastroenterol. 2019 Jan;53(1):e25–30.

22. Ganguly EK, Purvis L, Reynolds N, Akram S, Lidofsky SD, Zubarik R. An Accurate and Automated Method for Adenoma Detection Rate and Report Card Generation Utilizing Common Electronic Health Records. J Clin Gastroenterol. 2023 Aug 25;

23. Zand A, Sharma A, Stokes Z, Reynolds C, Montilla A, Sauk J, et al. An Exploration Into the Use of a Chatbot for Patients With Inflammatory Bowel Diseases: Retrospective Cohort Study. J Med Internet Res. 2020 May 26;22(5):e15589.

24. Li J, Wang X, Cai L, Sun J, Yang Z, Liu W, et al. An interpretable deep learning framework for predicting liver metastases in postoperative colorectal cancer patients using natural language processing and clinical data integration. Cancer Med. 2023 Sep;12(18):19337–51.

25. Atarere J, Naqvi H, Haas C, Adewunmi C, Bandaru S, Allamneni R, et al. Applicability of Online Chat-Based Artificial Intelligence Models to Colorectal Cancer Screening. Dig Dis Sci. 2024 Mar;69(3):791–7.

26. Laique SN, Hayat U, Sarvepalli S, Vaughn B, Ibrahim M, McMichael J, et al. Application of optical character recognition with natural language processing for large-scale quality metric data extraction in colonoscopy reports. Gastrointest Endosc. 2021 Mar;93(3):750–7.

27. Mehrotra A, Dellon ES, Schoen RE, Saul M, Bishehsari F, Farmer C, et al. Applying a natural language processing tool to electronic health records to assess performance on colonoscopy quality measures. Gastrointest Endosc. 2012 Jun;75(6):1233–1239.e14.

28. Pradhan F, Fiedler A, Samson K, Olivera-Martinez M, Manatsathit W, Peeraphatdit T. Artificial intelligence compared with human-derived patient educational materials on cirrhosis. Hepatol Commun. 2024 Mar 1;8(3):e0367.

29. Yeo YH, Samaan JS, Ng WH, Ting PS, Trivedi H, Vipani A, et al. Assessing the performance of ChatGPT in answering questions regarding cirrhosis and hepatocellular carcinoma. Clin Mol Hepatol. 2023 Jul;29(3):721–32.

30. Van Vleck TT, Chan L, Coca SG, Craven CK, Do R, Ellis SB, et al. Augmented intelligence with natural language processing applied to electronic health records for identifying patients with non-alcoholic fatty liver disease at risk for disease progression. Int J Med Inf. 2019 Sep;129:334–41.

31. Hou JK, Chang M, Nguyen T, Kramer JR, Richardson P, Sansgiry S, et al. Automated identification of surveillance colonoscopy in inflammatory bowel disease using natural language processing. Dig Dis Sci. 2013 Apr;58(4):936–41.

32. Gravina AG, Pellegrino R, Palladino G, Imperio G, Ventura A, Federico A. Charting new AI education in gastroenterology: Cross-sectional evaluation of ChatGPT and perplexity AI in medical residency exam. Dig Liver Dis Off J Ital Soc Gastroenterol Ital Assoc Study Liver. 2024 Mar 18;S1590-8658(24)00302-5.

33. Lim DYZ, Tan YB, Koh JTE, Tung JYM, Sng GGR, Tan DMY, et al. ChatGPT on guidelines: Providing contextual knowledge to GPT allows it to provide advice on appropriate colonoscopy intervals. J Gastroenterol Hepatol. 2024 Jan;39(1):81–106.

34. Samaan JS, Yeo YH, Ng WH, Ting PS, Trivedi H, Vipani A, et al. ChatGPT’s ability to comprehend and answer cirrhosis related questions in Arabic. Arab J Gastroenterol Off Publ Pan-Arab Assoc Gastroenterol. 2023 Aug;24(3):145–8.

35. Wagholikar K, Sohn S, Wu S, Kaggal V, Buehler S, Greenes R, et al. Clinical Decision Support for Colonoscopy Surveillance Using Natural Language Processing. In: 2012 IEEE Second International Conference on Healthcare Informatics, Imaging and Systems Biology [Internet]. 2012 [cited 2024 May 1]. p. 12–21. Available from: https://ieeexplore.ieee.org/document/6366186

36. Imler TD, Morea J, Imperiale TF. Clinical decision support with natural language processing facilitates determination of colonoscopy surveillance intervals. Clin Gastroenterol Hepatol Off Clin Pract J Am Gastroenterol Assoc. 2014 Jul;12(7):1130–6.

37. Pereyra L, Schlottmann F, Steinberg L, Lasa J. Colorectal Cancer Prevention: Is Chat Generative Pretrained Transformer (Chat GPT) ready to Assist Physicians in Determining Appropriate Screening and Surveillance Recommendations? J Clin Gastroenterol. 2024 Feb 7;

38. Kong Q, Ju K, Wan M, Liu J, Wu X, Li Y, et al. Comparative analysis of large language models in medical counseling: A focus on *Helicobacter pylori* infection. Helicobacter. 2024 Jan;29(1):e13055.

39. Choo JM, Ryu HS, Kim JS, Cheong JY, Baek SJ, Kwak JM, et al. Conversational artificial intelligence (chatGPT^TM^) in the management of complex colorectal cancer patients: early experience. ANZ J Surg. 2024 Mar;94(3):356–61.

40. Seong D, Choi YH, Shin SY, Yi BK. Deep learning approach to detection of colonoscopic information from unstructured reports. BMC Med Inform Decis Mak. 2023 Feb 7;23(1):28.

41. Wang X, Xu X, Tong W, Liu Q, Liu Z. DeepCausality: A general AI-powered causal inference framework for free text: A case study of LiverTox. Front Artif Intell [Internet]. 2022 Dec 6 [cited 2024 May 1];5. Available from: https://www.frontiersin.org/articles/10.3389/frai.2022.999289

42. Harkema H, Chapman WW, Saul M, Dellon ES, Schoen RE, Mehrotra A. Developing a natural language processing application for measuring the quality of colonoscopy procedures. J Am Med Inform Assoc JAMIA. 2011 Dec;18 Suppl 1(Suppl 1):i150–156.

43. Ma H, Ma X, Yang C, Niu Q, Gao T, Liu C, et al. Development and evaluation of a program based on a generative pre-trained transformer model from a public natural language processing platform for efficiency enhancement in post-procedural quality control of esophageal endoscopic submucosal dissection. Surg Endosc. 2024 Mar;38(3):1264–72.

44. Huo B, McKechnie T, Ortenzi M, Lee Y, Antoniou S, Mayol J, et al. Dr. GPT will see you now: the ability of large language model-linked chatbots to provide colorectal cancer screening recommendations. Health Technol [Internet]. 2024 Mar 4 [cited 2024 Apr 30]; Available from: 10.1007/s12553-024-00836-9

45. Peng W, Feng Y, Yao C, Zhang S, Zhuo H, Qiu T, et al. Evaluating AI in medicine: a comparative analysis of expert and ChatGPT responses to colorectal cancer questions. Sci Rep. 2024 Feb 3;14(1):2840.

46. Lahat A, Shachar E, Avidan B, Shatz Z, Glicksberg BS, Klang E. Evaluating the use of large language model in identifying top research questions in gastroenterology. Sci Rep. 2023 Mar 13;13(1):4164.

47. Lahat A, Shachar E, Avidan B, Glicksberg B, Klang E. Evaluating the Utility of a Large Language Model in Answering Common Patients’ Gastrointestinal Health-Related Questions: Are We There Yet? Diagnostics. 2023 Jan;13(11):1950.

48. Zhou J, Li T, Fong SJ, Dey N, González-Crespo R. Exploring ChatGPT’s Potential for Consultation, Recommendations and Report Diagnosis: Gastric Cancer and Gastroscopy Reports’ Case. Int J Interact Multimed Artif Intell. 2023;8(Regular Issue):7–13.

49. Truhn D, Loeffler CM, Müller-Franzes G, Nebelung S, Hewitt KJ, Brandner S, et al. Extracting structured information from unstructured histopathology reports using generative pre-trained transformer 4 (GPT-4). J Pathol. 2024 Mar;262(3):310–9.

50. Denny JC, Peterson JF, Choma NN, Xu H, Miller RA, Bastarache L, et al. Extracting timing and status descriptors for colonoscopy testing from electronic medical records. J Am Med Inform Assoc JAMIA. 2010;17(4):383–8.

51. Gorelik Y, Ghersin I, Maza I, Klein A. Harnessing language models for streamlined postcolonoscopy patient management: a novel approach. Gastrointest Endosc. 2023 Oct;98(4):639–641.e4.

52. Stidham RW, Yu D, Zhao X, Bishu S, Rice M, Bourque C, et al. Identifying the Presence, Activity, and Status of Extraintestinal Manifestations of Inflammatory Bowel Disease Using Natural Language Processing of Clinical Notes. Inflamm Bowel Dis. 2023 Apr 3;29(4):503–10.

53. Ananthakrishnan AN, Cai T, Savova G, Cheng SC, Chen P, Perez RG, et al. Improving case definition of Crohn’s disease and ulcerative colitis in electronic medical records using natural language processing: a novel informatics approach. Inflamm Bowel Dis. 2013 Jun;19(7):1411–20.

54. Schneider CV, Li T, Zhang D, Mezina AI, Rattan P, Huang H, et al. Large-scale identification of undiagnosed hepatic steatosis using natural language processing. eClinicalMedicine. 2023 Aug;62:102149.

55. Benson R, Winterton C, Winn M, Krick B, Liu M, Abu-el-rub N, et al. Leveraging Natural Language Processing to Extract Features of Colorectal Polyps From Pathology Reports for Epidemiologic Study. JCO Clin Cancer Inform [Internet]. 2023 [cited 2024 May 1];7. Available from: https://www.ncbi.nlm.nih.gov/pmc/articles/PMC10166420/

56. Benedicenti F, Pessarelli T, Corradi M, Michelon M, Nandi N, Lampertico P, et al. Mirror, mirror on the wall, who is the best of them all? Artificial intelligence versus gastroenterologists in solving clinical problems. Gastroenterol Rep. 2023 Jan 1;11:goad052.

57. Nayor J, Borges LF, Goryachev S, Gainer VS, Saltzman JR. Natural Language Processing Accurately Calculates Adenoma and Sessile Serrated Polyp Detection Rates. Dig Dis Sci. 2018 Jul;63(7):1794–800.

58. Imler TD, Morea J, Kahi C, Imperiale TF. Natural language processing accurately categorizes findings from colonoscopy and pathology reports. Clin Gastroenterol Hepatol Off Clin Pract J Am Gastroenterol Assoc. 2013 Jun;11(6):689– 94.

59. Raju GS, Lum PJ, Slack RS, Thirumurthi S, Lynch PM, Miller E, et al. Natural language processing as an alternative to manual reporting of colonoscopy quality metrics. Gastrointest Endosc. 2015 Sep;82(3):512–9.

60. Bae JH, Han HW, Yang SY, Song G, Sa S, Chung GE, et al. Natural Language Processing for Assessing Quality Indicators in Free-Text Colonoscopy and Pathology Reports: Development and Usability Study. JMIR Med Inform. 2022 Apr 15;10(4):e35257.

61. Song G, Chung SJ, Seo JY, Yang SY, Jin EH, Chung GE, et al. Natural Language Processing for Information Extraction of Gastric Diseases and Its Application in Large-Scale Clinical Research. J Clin Med. 2022 May 24;11(11):2967.

62. Li D, Udaltsova N, Layefsky E, Doan C, Corley DA. Natural Language Processing for the Accurate Identification of Colorectal Cancer Mismatch Repair Status in Lynch Syndrome Screening. Clin Gastroenterol Hepatol Off Clin Pract J Am Gastroenterol Assoc. 2021 Mar;19(3):610–612.e1.

63. Denny JC, Choma NN, Peterson JF, Miller RA, Bastarache L, Li M, et al. Natural language processing improves identification of colorectal cancer testing in the electronic medical record. Med Decis Mak Int J Soc Med Decis Mak. 2012;32(1):188–97.

64. Becker M, Kasper S, Böckmann B, Jöckel KH, Virchow I. Natural language processing of German clinical colorectal cancer notes for guideline-based treatment evaluation. Int J Med Inf. 2019 Jul;127:141–6.

65. Blumenthal DM, Singal G, Mangla SS, Macklin EA, Chung DC. Predicting Non-Adherence with Outpatient Colonoscopy Using a Novel Electronic Tool that Measures Prior Non-Adherence. J Gen Intern Med. 2015 Jun;30(6):724–31.

66. Imler TD, Sherman S, Imperiale TF, Xu H, Ouyang F, Beesley C, et al. Provider-specific quality measurement for ERCP using natural language processing. Gastrointest Endosc. 2018 Jan;87(1):164–173.e2.

67. Cankurtaran RE, Polat YH, Aydemir NG, Umay E, Yurekli OT. Reliability and Usefulness of ChatGPT for Inflammatory Bowel Diseases: An Analysis for Patients and Healthcare Professionals. Cureus. 15(10):e46736.

68. Rammohan R, Joy MV, Magam SG, Natt D, Magam SR, Pannikodu L, et al. Understanding the Landscape: The Emergence of Artificial Intelligence (AI), ChatGPT, and Google Bard in Gastroenterology. Cureus [Internet]. 2024 Jan 8 [cited 2024 Apr 30];16(1). Available from: https://www.cureus.com/articles/219958-understanding-the-landscape-the-emergence-of-artificial-intelligence-ai-chatgpt-and-google-bard-in-gastroenterology

69. Nguyen Wenker T, Natarajan Y, Caskey K, Novoa F, Mansour N, Pham HA, et al. Using Natural Language Processing to Automatically Identify Dysplasia in Pathology Reports for Patients With Barrett’s Esophagus. Clin Gastroenterol Hepatol Off Clin Pract J Am Gastroenterol Assoc. 2023 May;21(5):1198–204.

70. Tinmouth J, Swain D, Chorneyko K, Lee V, Bowes B, Li Y, et al. Validation of a natural language processing algorithm to identify adenomas and measure adenoma detection rates across a health system: a population-level study. Gastrointest Endosc. 2023 Jan;97(1):121–129.e1.

71. Sada Y, Hou J, Richardson P, El-Serag H, Davila J. Validation of Case Finding Algorithms for Hepatocellular Cancer From Administrative Data and Electronic Health Records Using Natural Language Processing. Med Care. 2016 Feb;54(2):e9–14.

72. Wang Y, Huang Y, Nimma IR, Pang S, Pang M, Cui T, et al. Validation of GPT-4 for clinical event classification: A comparative analysis with ICD codes and human reviewers. J Gastroenterol Hepatol. 2024 Apr 16;

73. Mandrekar JN. Measures of Interrater Agreement. J Thorac Oncol. 2011 Jan 1;6(1):6–7.

74. Omar M, Brin D, Glicksberg B, Klang E. Utilizing Natural Language Processing and Large Language Models in the Diagnosis and Prediction of Infectious Diseases: A Systematic Review. Am J Infect Control [Internet]. 2024 Apr 5 [cited 2024 Apr 22];0(0). Available from: https://www.ajicjournal.org/article/S0196-6553(24)00159-7/abstract

75. Casey A, Davidson E, Poon M, Dong H, Duma D, Grivas A, et al. A systematic review of natural language processing applied to radiology reports. BMC Med Inform Decis Mak. 2021 Jun 3;21:179.

76. Cheng S, Chang C, Chang W, Wang H, Liang C, Kishimoto T, et al. The now and future of ChatGPT and GPT in psychiatry. Psychiatry Clin Neurosci. 2023 Nov;77(11):592–6.

77. Oh N, Choi GS, Lee WY. ChatGPT goes to the operating room: evaluating GPT-4 performance and its potential in surgical education and training in the era of large language models. Ann Surg Treat Res. 2023 May;104(5):269–73.

78. Wang J, Deng H, Liu B, Hu A, Liang J, Fan L, et al. Systematic Evaluation of Research Progress on Natural Language Processing in Medicine Over the Past 20 Years: Bibliometric Study on PubMed. J Med Internet Res. 2020 Jan 23;22(1):e16816.

79. Poon AIF, Sung JJY. Opening the black box of AI-Medicine. J Gastroenterol Hepatol. 2021 Mar;36(3):581–4.

80. Herington J, McCradden MD, Creel K, Boellaard R, Jones EC, Jha AK, et al. Ethical Considerations for Artificial Intelligence in Medical Imaging: Deployment and Governance. J Nucl Med Off Publ Soc Nucl Med. 2023 Oct;64(10):1509–15.

