## Supplementary material for "Emerging Applications of NLP and Large Language Models in Gastroenterology and Hepatology: A Systematic Review": Table 1

**Table 1: Summary of the included studies.**

| author | Year | Data Type + Sample Size | Model | Model Task | Main Result |
|---|---|---|---|---|---|
| | | | **Gastroenterology** | | |
| **Kong et al.** | 2024 | 15 questions related to H. pylori | ChatGPT 4.0, ChatGPT 3.5, ERNIE Bot 4.0 | Counseling on H. pylori infection | Satisfactory accuracy and comprehensibility; low completeness |
| **Lahat et al.** | 2023 | 110 real-life patient questions | GPT | Answering patient questions | Varied performance in accuracy and clarity |
| **Truhn et al.** | 2024 | 100 colorectal cancer reports | GPT-4 | Extracting structured information | High accuracy in extraction of cancer stages |
| **Zhou et al.** | 2023 | NR | GPT-3.5 and GPT-4 | Medical consultation and report analysis | High effectiveness in consultation accuracy |
| **Choo et al.** | 2024 | 30 patients with advanced colorectal cancer | GPT | Formulating management plans | High concordance with multidisciplinary team plans |
| **Huo et al.** | 2024 | Responses for nine patient cases | ChatGPT, Bing Chat, Google Bard, Claude 2 | Providing screening recommendations | Variable advice quality, some aligned with clinical guidelines |
| **Imler et al.** | 2013 | 500 colonoscopy and pathology reports | cTAKES NLP engine | Categorizing pathology findings | High accuracy in identifying pathology levels |

| | | | | | |
|---|---|---|---|---|---|
| **Lim et al.** | 2024 | 62 example case scenarios, tested three times | GPT-4 | Providing advice on colonoscopy intervals | Consistent high adherence to guidelines |
| **Imler et al.** | 2014 | 10,798 colonoscopy reports, 6,379 linked to pathology | Clinical text analysis and knowledge extraction system (cTAKES) | Determining colonoscopy surveillance intervals | High agreement with standards, improved guideline adherence |
| **Bae et al.** | 2022 | 2,425 colonoscopy and pathology reports | Regular expressions and smartTA | Assessing quality indicators | High accuracy and matching expert review |
| **Denny et al.** | 2012 | 200 patients | KnowledgeMap Concept Identifier | Identifying colorectal cancer tests in EMRs | Superior recall compared to manual and billing records |
| **Lahat et al.** | 2023 | 20 research questions | GPT | Generating gastroenterology research questions | Relevant and clear questions generated, low originality |
| **Laique et al.** | 2021 | 35,914 colonoscopy reports | Optical Character Recognition (OCR) and NLP | Extracting quality metrics | High accuracy in detecting clinical variables |
| **Blumenthal et al.** | 2015 | 1,531 patients | NLP tool called QPID | Predicting non-adherence to colonoscopy | Effective prediction supported by Non-Adherence Ratio |
| **Harkema et al.** | 2011 | 679 colonoscopy and pathology reports | Rule-based NLP engine | Quality measurement in colonoscopy | High accuracy in automated quality assessment |

| | | | | | |
|---|---|---|---|---|---|
| **Raju et al.** | 2015 | 12,748 colonoscopy patients | Custom NLP software | Reporting colonoscopy quality metrics | Comparable to manual methods, high ADR detection |
| **Nayor et al.** | 2018 | 8,032 screening colonoscopies | NLP pipeline | Calculating adenoma and serrated polyp detection rates | Perfect precision and recall |
| **Atarere et al.** | 2024 | 20 questions using AI models | ChatGPT, BingChat, and YouChat™ | CRC screening advice | High inter-rater reliability, variable accuracy |
| **Seong et al.** | 2023 | 280,668 colonoscopy reports | LSTM, BioBERT, Bi-LSTM-CRF | Extracting information from reports | Superior performance in extracting colonoscopic findings |
| **Lee et al.** | 2019 | 800 colonoscopy reports | Commercial NLP tool | Identifying quality and large polyps | High sensitivity and specificity in data extraction |
| **Denny et al.** | 2010 | 200 patients | KnowledgeMap concept identifier | Detecting colonoscopy timing and status | High recall and precision, improved scheduling |
| **Parthasarathy et al.** | 2020 | 323,494 colonoscopy patients | NLP | Diagnosing serrated polyposis syndrome | High accuracy in identifying SPS, improved detection rates |
| **Rammohan et al.** | 2024 | NR | GPT-4 and Bard | Answering standard gastroenterology questions | ChatGPT 4.0 more reliable and accurate than Bard |
| **Pereyra et al.** | 2024 | 238 physicians | GPT-3.5 | Assessing CRC screening recommendations | Lower performance compared to physicians, variable responses |
| **Song et al.** | 2022 | 1,000 validation, 248,966 | Custom NLP pipeline | Extracting information from EGD reports | High sensitivity, precision, and accuracy in data extraction |

| | | | | | |
|---|---|---|---|---|---|
| | | application EGD reports | | | |
| **Peng et al.** | 2024 | 131 colorectal cancer questions | GPT-3.5 | Answering CRC-related questions | High reproducibility, less comprehensive than expert answers |
| **Tinmouth et al.** | 2023 | 1,450 pathology reports | NLP | Identifying adenomas for ADR | High accuracy, supported system-level ADR measurement |
| **Mehrotra et al.** | 2012 | 24,157 colonoscopy reports | NLP (C-QUAL) | Assessing colonoscopy quality measures | Identified variability in provider performance |
| **Becker et al.** | 2019 | 2,513 German clinical notes from 500 patients | German-specific NLP pipeline | Guideline-based treatment evaluation | High precision and recall in extracting treatment information |
| **Hou et al.** | 2013 | 575 colonoscopy pathology reports | Automated Retrieval Console (ARC) | Identifying surveillance colonoscopy | Effective in classifying surveillance colonoscopy |
| **Gorelik et al.** | 2023 | 20 clinical scenarios | GPT-4 | Post-colonoscopy patient management | High compliance with guidelines, accurate recommendations |
| **Samaan et al.** | 2023 | 91 questions on liver cirrhosis | GPT | Answering cirrhosis-related questions | Lower accuracy in Arabic, significant discrepancies |
| **Cankurtaran et al.** | 2023 | 20 questions on Crohn's disease and ulcerative colitis | GPT | Responding to IBD queries | Higher reliability in professional context, variable in patients |

| | | | | | |
|---|---|---|---|---|---|
| **Wenker et al.** | 2023 | 1,000 patients for NLP validation | CLAMP NLP software | Identifying dysplasia in Barrett's Esophagus | High accuracy, sensitivity, and precision in dysplasia identification |
| **Imler et al.** | 2018 | 23,674 ERCP procedures | NLP | Quality measurement for ERCP | High accuracy and precision in identifying quality measures |
| **Li et al.** | 2021 | 5,570 patients | NLP | Identifying Lynch Syndrome for MMR screening | High accuracy in classifying MMR IHC results |
| **Li et al.** | 2022 | 22,206 patients across various tests | ENDOANGEL-AS NLP and deep learning | Identifying high-risk patients for surveillance | High accuracy in patient identification and risk classification |
| **Wagholikar et al.** | 2012 | 53 patients | NLP | Providing colonoscopy surveillance guidance | High effectiveness in clinical decision support |
| **Sciberras et al.** | 2024 | 38 questions from IBD patients | GPT-3.5 | Generating responses to IBD patient queries | High accuracy and moderate completeness |
| **Stidham et al.** | 2023 | 1,240 patients with IBD | NLP | Detecting and inferring EIM activity status | High effectiveness in improving disease management |
| **Ganguly et al.** | 2023 | 2,276 colonoscopy procedures | NLP | Adenoma detection and report card generation | High sensitivity, specificity, and consistency |
| **Ma et al.** | 2024 | 165 esophageal ESD cases | GPT-3.5 | Post-procedural quality control for esophageal ESD | Improved efficiency and accuracy in quality assessments |
| **Gravina et al.** | 2024 | Questions from 2023 Italian medical exam | GPT 3.5 and Perplexity AI | Answering medical residency exam questions | High accuracy in exam responses, educational potential |
| **Fevrier et al.** | 2020 | 401,566 colonoscopy linked | SAS® PERL NLP tool | Extracting data from colonoscopy reports | High performance in classifying key clinical variables |

| | | | | | |
|---|---|---|---|---|---|
| | | with pathology reports | | | |
| **Benson et al.** | 2023 | 24,584 pathology reports | NLP pipeline | Extracting features of colorectal polyps | High precision and efficiency in feature extraction |
| **Zand et al.** | 2020 | 16,453 lines of dialog from 424 patients | NLP model | Developing a chatbot for IBD patient support | Effective in categorizing patient dialog |
| **Ananthakrishnan et al.** | 2013 | 1,200 patients for Crohn's and UC | NLP techniques | Improving EMR case definitions for IBD | Enhanced sensitivity and predictive value in identifying IBD |
| **Wang et al.** | 2024 | 200 medical discharge summaries | GPT-4 | Classifying GI bleeding events | High accuracy, outperforms ICD coding |
| | | | **Hepatology** | | |
| **Benedicenti et al.** | 2023 | 56 gastroenterologists, 25 residents, 31 specialists | GPT-3 | Answering clinical vignettes on Hepatology and Gastroenterology | Demonstrated improvement over time, underperformed vs. humans |
| **Li et al.** | 2023 | 1,463 postoperative colorectal cancer patients | NLP and machine learning integration | Predicting liver metastases | High accuracy in risk prediction |
| **Wang et al.** | 2022 | LiverTox database | DeepCausality framework | Causal inference for drug-induced liver injury | High accuracy and concordance with clinical guidelines |
| **Yeo et al.** | 2023 | 164 questions about cirrhosis and | GPT | Providing answers on cirrhosis and HCC | Good accuracy, but not comprehensive |

| | | | | | |
|---|---|---|---|---|---|
| | | hepatocellular carcinoma | | | |
| **Sherman et al.** | 2024 | 3,134 patients with liver disease | NLP | Classifying liver disease pathology | High predictive values, enabled insights into disease progression |
| **Van Vleck et al.** | 2019 | 38,575 patients | CLiX clinical NLP engine | Identifying NAFLD patients and disease progression | High sensitivity in tracking disease progression |
| **Sada et al.** | 2016 | 1,138 patients identified from ICD-9 codes | Automated Retrieval Console (ARC) | Improving identification of hepatocellular cancer | Enhanced accuracy in HCC case identification |
| **Pradhan et al.** | 2024 | 22 patients/caregivers and transplant hepatologists | Multiple LLMs | Generating patient educational materials about cirrhosis | Comparable understandability and accuracy to human materials |
| **Schneider et al.** | 2023 | 2.15 million pathology and 2.7 million imaging reports | Rule-based NLP algorithm | Identifying hepatic steatosis | Effective detection of undiagnosed cases |
